# Discovering Digital Biomarkers of Panic Attack Risk in Consumer Wearables Data

**DOI:** 10.1101/2023.03.01.23286647

**Authors:** Ellen W. McGinnis, Shania Lunna, Isabel Berman, Bryn C. Loftness, Skylar Bagdon, Christopher M. Danforth, Matthew Price, William E. Copeland, Ryan S. McGinnis

## Abstract

Panic attacks are an impairing mental health problem that impacts more than one out of every 10 adults in the United States (US). Clinical guidelines suggest panic attacks occur without warning and their unexpected nature worsens their impact on quality of life. Individuals who experience panic attacks would benefit from advance warning of when an attack is likely to occur so that appropriate steps could be taken to manage or prevent it. Our recent work suggests that an individual’s likelihood of experiencing a panic attack can be predicted by self-reported mood and community-level Twitter-derived mood the previous day. Prior work also suggests that physiological markers may indicate a pending panic attack. However, the ability of objective physiological, behavioral, and environmental measures to predict next-day panic attacks has not yet been explored. To address this question, we consider data from 38 individuals who regularly experienced panic attacks recruited from across the US. Participants responded to daily questions about their panic attacks for 28 days and provided access to data from their Apple Watches. Results indicate that objective measures of ambient noise (louder) and resting heart rate (higher) are related to the likelihood of experiencing a panic attack the next day. These preliminary results suggest, for the first time, that panic attacks may be predictable from data passively collected by consumer wearable devices, opening the door to improvements in how panic attacks are managed and to the development of new preventative interventions.

**Clinical Relevance:** Objective data from consumer wearables may predict when an individual is at high risk for experiencing a next-day panic attack. This information could guide treatment decisions, help individuals manage their panic, and inform the development of new preventative interventions.

## I. Introduction

More than one out of every four US adults have experienced a panic attack in their lifetime with over one in 10 experiencing at least one in the last year [1]. Physiological symptoms of panic attacks include increased heart rate, hyperventilation, shaking, and feelings of faintness, and are paired with psychological symptoms including a self-perceived lack of control, and fear of experiencing another attack [2]. Clinical guidelines suggest panic attacks occur without warning or a triggering event [2]. However, our previous work demonstrates 98% of our 85 participants who experienced regular panic attacks could identify a trigger of their latest attack [3]. Health was the most common trigger (∼40%, the study co-occurred with the start of COVID-19 lockdowns in the US), followed by conflict, performance, and workload. Only two participants reported being unsure of what triggered their latest attack [3]. Given the prevalence of self-reported triggers in this study, it is important to further examine prospective factors that may lead to panic attacks and to investigate if it is possible to predict their occurrence.

Prospective studies on panic attacks are limited however, with those that do exist confirming associations with wellness behaviors such as caffeine consumption [4]) and identifying potential physiological markers of a pending attack (i.e., changes in heart rate 45 minutes prior to onset [5]). However, these studies are limited in size, have only considered individuals with panic disorder, and, use methods that limit generalizability and utility of the results (in-lab dosing of caffeine equivalent to more than 4 cups of coffee [4], requiring cumbersome sensor arrays [5]). Our prior work was the first to examine prospective markers of panic attacks in those without panic disorder and we found that an individual’s likelihood of experiencing an attack can be predicted by their self-reported mood and the Twitter-derived mood in their community the previous day[6]. Given the growing prevalence of consumer wearable devices that capture objective physiology, there is opportunity to explore within-person changes that occur prior to the onset of an attack that could serve as prospective biomarkers of panic attack risk.

Prior work also suggests external factors, such as community mood [6], or a global pandemic [3], may be associated with panic. Similarly, more abstract external stimuli like ambient noise is known to increase stress levels and may lead to atypical activation of the autonomic nervous system (e.g., [7]). This connection may be particularly salient for anxious individuals who attend more to potential external threats [8]. Recent advances in consumer wearable devices enable objective measurement of external stimuli such as ambient noise. However, it is not yet known how these measurements may relate to panic attacks.

We herein examine the ability of objective measures of physiology and external stimuli from consumer wearable devices to prospectively predict next-day panic attack risk. Results could ultimately inform future prevention efforts.

## II. Methods

### A. Fully Remote Experimental Protocol

Facebook ads targeting users identified as “highly anxious” and having “interest in Apple Watches” recruited participants remotely over 72 hours in Spring 2022. Eligible participants had to 1) live in the United States, 2) own an iPhone and 3) an Apple Watch, 4) be at least 18 years old, and 5) have experienced at least one panic attack in the last week. The University of Vermont Institutional Review Board approved the study protocol. After e-consenting, participants were asked to complete a survey including items on demographic information, mental health history, and panic attack history. For the 28 days thereafter, participants complete a daily survey including items on if they had experienced a panic attack and associated descriptive information. Participants were asked to wear their Apple Watch every day and upload their data weekly. Participants earned a $20 Amazon gift card each week they were enrolled in the study.

### B. Characteristics of the Sample

There was significant attrition in this completely remote study. To start, only 87 of 107 participants who signed the consent form completed at least one daily survey. At the end of each week, participants were withdrawn by the study team if they had completed less than 15% of their surveys. Thirty-five participants were withdrawn after week one, 25 after week two, and one after week three. Thirty-eight participants uploaded any Apple Watch data appropriately. Two participants uploaded photos of themselves wearing their Apple Watches, and the rest of the participants did not attempt to upload any data. Four of the 38 participants who uploaded data did so at the end of week one only, two participants uploaded data at the end of week two, and 32 participants uploaded their Apple Watch data at the end of week four.

Participants with Apple Watch data uploaded were 79% female, 33% minority race/ethnicity, ranged in age from 18-69 (M: 38.92, SD: 13.36), lived across 7 US states, earned an average of $68,000 annually (SD: $25,000) and 78% had a bachelor’s degree or more. Half (50%) reporting having a mental health diagnosis, and an average of 5.56 (SD: 9.23) panic attacks in the month prior to study participation.

Attrition analyses revealed that Apple Watch uploaders did not significantly differ from those who did not upload data on educational attainment, annual income, age, gender, race/ethnicity, mental health diagnostic status, or number of panic attacks. All participants who uploaded any Apple Watch data were included in analyses.

### C. Apple Watch Metrics of Physiology and External Stimuli

Features extracted from the Apple Watch were daily measures of resting heart rate (RHR), heart rate variability (HRV), respiratory rate (RR), and ambient noise levels (Noise). The watch also reports estimates of distance walked, steps taken, and stair flights climbed. We combined these metrics into a single aggregate activity feature (Activity) by averaging subject-specific z-scores of each metric for each participant each day. Summary statistics and sample sizes for watch-derived features are reported in Table I. Not all measures were available from every participant every day. To account for expected subject-specific baselines in physiological parameters, RHR, HRV, and RR measures from each subject were adjusted by their associated mean over the study period prior to performing the analysis described next.

**TABLE I.** Characteristics of Apple Watch Data

| Measures | People | Days | Panic Days | Mean (SD) |
| --- | --- | --- | --- | --- |
| HRV | 31 | 273 | 48 | 41.41 (18.54) |
| RHR | 31 | 241 | 40 | 66.09 (7.50) |
| Activity<br>Distance<br>Flights<br>Steps | 31 | 264 | 44 | 2.77 (1.69)<br>4.36 (3.22)<br>5,934 (3,621) |
| RR | 18 | 135 | 32 | 16.48 (1.72) |
| Noise | 10 | 125 | 23 | 62.51 (5.43) |

HRV = Heart Rate Variability in milliseconds, RHR = Resting Heart Rate in beats per minute, Flights = Stair flights climbed, Distance = Distance walked in miles, RR = Respiratory Rate in breaths per minute, Noise = Ambient Noise Level in decibels, People = Number of participants with the associated measure, Days = Number of days with measure available, Panic Days = Number of days with the measure available where panic attack occurred, SD = standard deviation. Means winzorized at 95%.

### D. Analytical Plan

Independence of Apple Watch-derived features are assessed using Pearson correlation where each day of data from each subject is considered a unique observation. While this approach yields overly optimistic power, it also helps expose any potential relationships between features.

Mixed Regressions, with an autoregressive covariance structure were used to estimate the prevalence of a next-day panic attack. In this approach, participant ID is introduced as a cluster (class) variable to account for repeated, correlated observations within individuals. Robust variance estimates (i.e., sandwich type estimates) adjusted the standard errors of the parameter estimates for the within-person nesting of observations. The alpha value for significance testing was set at <.05 and interpreted as trend level at <.01. All statistical analyses were performed in SPSS (Version 28.0.1.1, IBM). Individual-level watch-derived measures were examined using univariate models for outcomes of next-day panic attack occurrence using lag sequential analyses. In each model, number of days since the start of the study, participant gender, race, and mental health diagnostic status were included as covariates as our prior work demonstrated significant associations with panic attack likelihood.

## III. Results

As indicated in Table II, correlation analysis reveals several significant, but weak relationships between the watch-derived features. Specifically, HRV deviation was negatively correlated with RHR and RR deviation, RHR deviation was positively correlated with RR deviation, and Noise was positively correlated with Activity and RR deviation. While significant, the strength of these associations suggests that the features within this watch-derived feature set are largely independent in this sample. The relationships are also in line with expected physiological relationships (e.g., negative relationship between HRV and RHR deviation, positive relationship between RHR and RR deviation) lending support to the measurements.

**TABLE II.** correlation between Apple Watch Features

| Measures | RHR | Activity | RR | Noise |
| --- | --- | --- | --- | --- |
| HRV | -.374* | -.056 | -.223* | -.072 |
| RHR | - | .076 | .114* | .127 |
| Activity | - | - | .074 | .134* |
| RR | - | - | - | .295* |

HRV = Heart Rate Variability deviation, RHR = Resting Heart Rate deviation, Activity = aggregate measure derived from distance walked, steps, and flights of stairs climbed, RR = Respiratory Rate deviation, Noise = Ambient Noise Level. The \* indicates statistical significance at the 0.05 level.

Controlling for number of days since the start of the study, participant gender, race, and mental health diagnostic status, univariate regression analysis reveals that increases in resting heart rate above an individual’s mean value during the study and higher ambient noise levels are associated with an increased likelihood of experiencing a panic attack the next day (Table III). Notably, we also explored ambient noise deviation and found a similarly significant linear effect (*b*=.003, S.E.= .009, CIs= .015-.052, *p* = .001).

**TABLE III.** Associations of Apple Watch Data with Next Day Panic Attack in Univariate Models

| Measures | Est. | Std. Error | CI's | p-value |
| --- | --- | --- | --- | --- |
| HRV | .001 | .003 | -.005 - .006 | .820 |
| RHR | .008 | .004 | -.001 - .017 | .064<sup>†</sup> |
| Activity | -.009 | .026 | -.061 - .042 | .729 |
| RR | -.010 | .039 | -.088 - .067 | .793 |
| Noise | .022 | .009 | .005 - .040 | .014* |

HRV = Heart Rate Variability deviation, RHR = Resting Heart Rate deviation, Activity = aggregate measure derived from distance walked, steps, and flights of stairs climbed, RR = Respiratory Rate deviation, Noise = Ambient Noise Level, CI's = confidence interval of the model estimate. The \* indicates statistical significance at the 0.05 level, <sup>†</sup> indicates trend-level significance.

Digging deeper into these results, we binned significant continuous metrics for better visualization. First, we see that small deviations in resting heart rate from a participant’s mean over the study results in large changes in their likelihood of experiencing a panic attack the next day (Fig. 1, top). Specifically, deviations as small as 1 BPM above, and 5 BPM below, the mean RHR can yield more than a 100% increase (5 BPM below: .09 vs. .19; 1 BPM above: .09 vs. .23) in likelihood for experiencing a next-day panic attack. This is manifested in data from an example subject (Fig. 1, bottom) where deviations of 3, 4 and 5 BPM above the mean correspond to a next-day panic attack.

**Figure 1.**
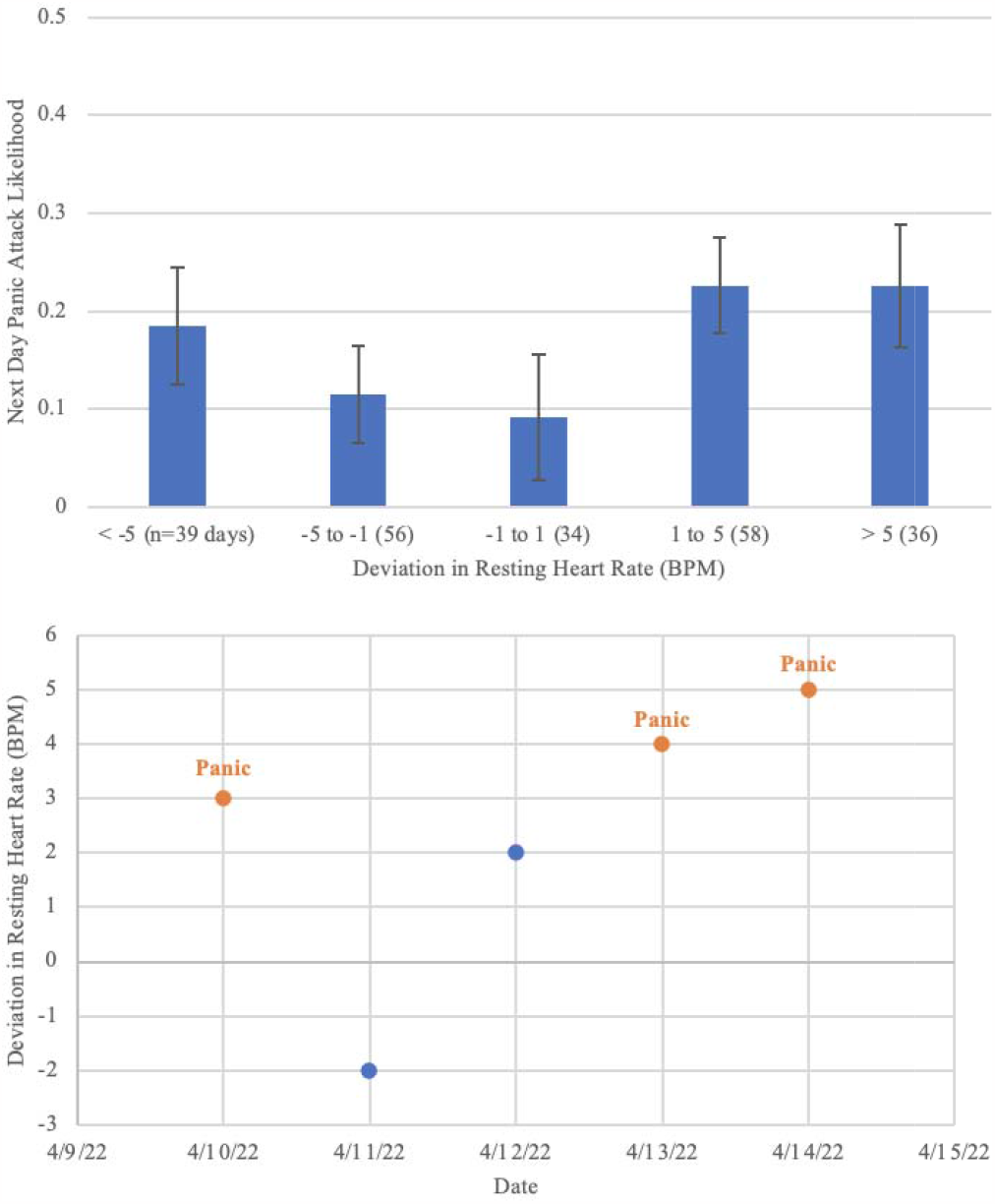
Larger deviations in resting heart rate above and below the mean are associated with a higher likelihood of experiencing a panic attack the next day (top). As seen in the case study of an example subject (bottom), deviations of 3, 4 and 5 beats per minute (BPM) correspond to subsequent days where a panic attack was experienced.

Similarly, relatively small changes in ambient noise level can yield dramatic changes in a participant’s likelihood of experiencing a panic attack the next day (Fig. 2, top). Specifically, for daily ambient noise levels on par with a typical conversation (∼60 dB), participants in this sample have a likelihood of experiencing a panic attack the next day of approximately .18. Increasing the ambient noise level above 70 dB increases the likelihood of experiencing a panic attack about 80% to .33. Interestingly, decreasing the ambient noise level below 57 dB dramatically reduces the likelihood of experiencing a panic attack (.18 to .01), but this may be impacted by a small sample size of quiet days (n=15). This manifests in data from an example subject (Fig. 2, bottom), where you can see that days with ambient noise above 65 dB are associated with subsequent panic attacks.

**Figure 2.**
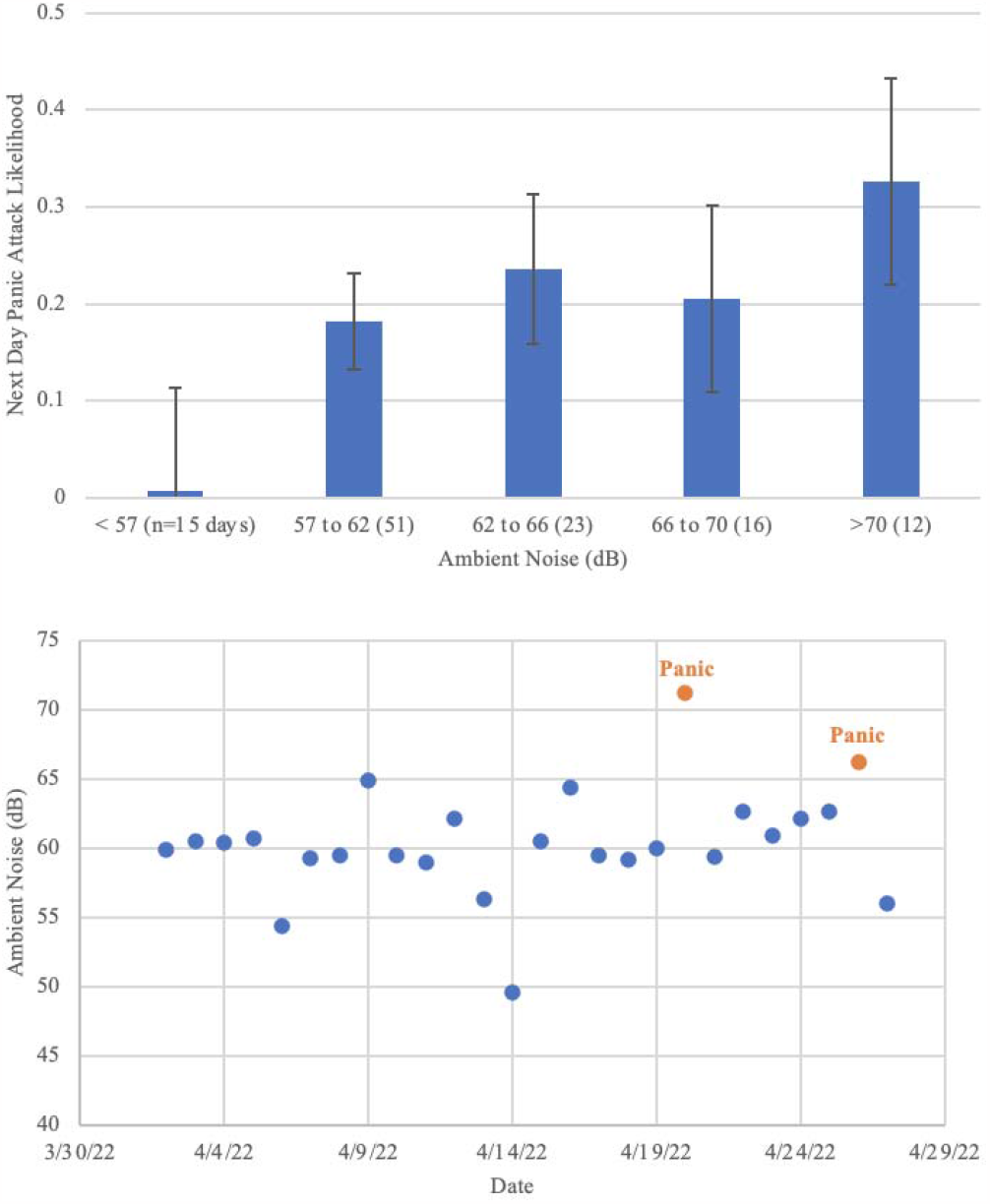
Higher ambient noise is associated with a higher likelihood of experiencing a panic attack the next day (top). As seen in the case study of an example subject (bottom), ambient noise above 65 dB corresponds to subsequent days where a panic attack was experienced.

## IV. Discussion

The purpose of this work was to investigate if quantitative prospective data measured by a popular consumer wearable device could be used to help predict when next-day panic attacks may occur. Mirroring our prior work, results suggest that both measures of a person’s internal physiological state (e.g., resting heart rate), and their external environment (e.g., ambient noise) are significant predictors of the likelihood of experiencing a panic attack within the next day.

These results fit well with what we know about how anxiety disorders impact the autonomic nervous system and how we respond to external stimuli. Systems responsible for allostasis (e.g., autonomic nervous system) allow the body to adapt to challenging situations while minimizing the amplitude of their response. Excess load from chronic stress, termed *allostatic load*, can be caused by frequent system activation, system over-reaction, and/or failure to regulate after stress [9]. Allostatic load is elevated in individuals with stress-related disorders such as anxiety and depression [10], [11]. Changes in resting heart rate are known to be induced by both sympathetic activation and parasympathetic withdrawal [12], characteristic of allostatic load, and have been linked to stress-related disorders (e.g., [13]). Similarly, those with anxiety and depression are known to be particularly sensitive to ambient noise which may lead to atypical activation of the autonomic nervous system (e.g., [7]). Our results indicating that elevated resting heart rate and ambient noise are associated with experiencing a next-day panic attack may imply elevated allostatic load may predispose an individual to an attack.

While preliminary, results suggest that it may be possible to predict panic attack risk prospectively using data from consumer wearable devices. This is significant considering clinical guidelines suggest panic attacks occur “out of the blue”. In the short term, these results could help individuals who experience panic attacks manage their panic by warning them when they are particularly susceptible. Similarly, this information could be a powerful addition to existing front-line psychoeducation interventions, providing critical data about how a panic attack occurs [14]. Longer-term, these results could inform the development of personalized, just-in-time interventions for panic attacks [3] that deliver a tailored therapy to participants right when they need it. In both cases, future work should consider machine learning based approaches for building predictive models of panic attack risk and specifically performance gains that may be achieved through model adaptation and personalization.

This study has several limitations. First, there was high attrition, particularly when considering the subset of participants with Apple Watch data. Second, most participants in the study identified as female and had completed some level of higher education. Attrition and demographics pose risks to the external validity of the study. Further research with a larger and more diverse sample are needed to confirm results and inform generalizable conclusions.

## V. Conclusion

We have presented promising preliminary data suggesting that we may be able to predict an individual’s risk for experiencing a panic attack based on objective data from a popular consumer wearable. While future work is needed to replicate these findings in a larger sample, this information could guide treatment decisions, help individuals manage their panic, and inform development of preventative interventions.

## Data Availability

All data produced in this manuscript are currently not available for request.

## Notes

* Research supported by the University of Vermont (UVM) through UVM Ventures and the National Institutes of Health (NIH Award MH123031)

### Competing Interest Statement

EWM and RSM are Co-founders of PanicMechanic, Inc., SB discloses stock ownership in PanicMechanic, Inc.

### Funding Statement

Research supported by the University of Vermont (UVM) through UVM Ventures and the National Institutes of Health (NIH Award MH123031).

### Author Declarations

University of Vermont IRB provided ethical approval for this work.

### Summary of Updates

Competing interest statement updated. All other files/details remain unchanged from the initial submission.

